# Glucose-loaded dopamine transporter is associated with mood altering effect of sweet foods and control over eating sweets

**DOI:** 10.1101/2024.04.15.24305879

**Authors:** Kyoungjune Pak, Jihyun Kim, Keunyoung Kim, Seongho Seo, Myung Jun Lee

## Abstract

Dopamine transporter (DAT) mediates reuptake of extracellular dopamine into presynaptic neurons. We investigated the effects of glucose loading on striatal DAT in healthy young adults who underwent ^18^F-FP-CIT positron emission tomography (PET) scans, and sweet taste questionnaire (STQ). Thirty-five healthy subjects were enrolled in this study. Each subject visited the institution three times, for three brain PET scans (two ^18^F-FP-CIT PET scans after the infusion of glucose or placebo and one ^18^F-Fluorodeoxyglucose PET scan). All subjects underwent STQ, 12-item self-reporting to evaluate subjects’ reactions to eating sweets, craving for sweets and degree of control over eating sweet foods (STQ 1: sensitivity to mood altering effect of sweets, and STQ 2: impaired control over eating sweet foods). We created Bayesian models separately with STQ 1, and STQ 2 as predictors, with DAT availability and brain glucose uptake as a dependent variable. From caudate, glucose-loaded DAT availability was significantly higher than placebo-loaded DAT availability, and from putamen, glucose-loaded DAT availability showed the higher trend than placebo-loaded DAT availability. STQ was positively associated with glucose-loaded DAT availability. The effect of STQ markedly overlapped with zero on placebo-loaded DAT availability, and brain glucose uptake. In conclusion, the change of striatal DAT availability after glucose loading is associated with the vulnerability to sweet foods. This may indicate that individuals with higher DAT availability after glucose loading experience a rapid clearance of synaptic dopamine after consuming sweet foods, potentially leading to a desire for additional sweet foods.

## INTRODUCTION

Obesity has nearly tripled worldwide since 1975 and has become one of the major public health threats. Obesity is a risk factor for malignancies of the colon [1], pancreas [2], thyroid [3], liver [4], and uterus [5] as well as for cardiovascular disease and diabetes mellitus [6].

The brain plays a critical role in controlling energy balance between energy intake and expenditure [7]. Food is a potent natural reinforcer, and fasting can further enhance its rewarding effects [8]. Dopamine is one of the neurotransmitters involved with eating behavior through modulation of the rewarding properties of food and the motivation and desire for food consumption [9]. Currently, there is no direct method to measure the synaptic dopamine concentration in the human brain. Therefore, positron emission tomography (PET) with dopamine receptor (DR) radiopharmaceuticals has been adopted to understand the role of dopaminergic system in obesity and eating behaviour. However, it is difficult to interpret the relationships between DR availability measured from PET scans and the synaptic dopamine level, which competes with DR radiopharmaceutical to bind DR [10,11]. In addition, brain DR availability is not different between lean and overweight/obese subjects from a recent meta-analysis [12].

Until recently, dopamine transporter (DAT), mediating reuptake of extracellular dopamine into presynaptic neurons [13], has not received as much attention in the area of eating behaviour, as DAT availability from brain PET scan was not correlated with body mass index (BMI) in previous studies [14-16]. However, the change of striatal DAT after glucose loading was reported by our group for the first time in humans [17]. Insulin has been shown to contribute to striatal DAT function by increasing DAT recruitment to the plasma membrane, and enhancing dopamine clearance via phosphatidylinositol 3-kinase signaling pathway, and downstream activation of Akt [13].

To address the effects of glucose loading on striatal DAT, we analyzed healthy young adults (mean 24.4 years) who underwent ^18^F-FP-CIT PET scans, and sweet taste questionnaire (STQ), which is a tool used to assess individual preferences and responses to sweet foods. In addition, ^18^F-FP-FDG PET scans were analyzed to investigate the association of STQ with striatal glucose metabolism. We used Bayesian hierarchical modeling to estimate the effects of 1) sensitivity to mood altering effect of sweet foods, and 2) impaired control over eating sweets on striatal DAT and glucose metabolism.

## METHODS

### Subjects

All participants signed an informed consent form prior to participation. Thirty-five healthy, male subjects were enrolled in this study. Subjects who had more than 10% change in weight over six months, were heavy smokers, or gave history of drug abuse, brain injury, neuropsychological disorders, or endocrine disorders were excluded. On the day of each visit, the subjects were instructed to fast overnight for at least 12 hours and abstain from smoking and alcohol consumption. The subjects visited the institution between 11 am and 12 pm to avoid the effect of diurnal variations in dopamine. During the visits, the height (m) and weight (kg) of the subject were measured. The body mass index (BMI) was calculated as follows: weight/height^-2^. The participants in this study were included in a previous study of striatal DAT changes after glucose loading [17,18]. This study was approved by the institutional review board of Pusan National University Hospital (PNUH-1707-019-057).

### Image acquisition

Each subject visited the institution three times, on separate days, for three brain PET scans (two ^18^F-FP-CIT PET scans and one ^18^F-FDG PET scan). During the visits, the height (m) and weight (kg) of the subject were measured. For ^18^F-FP-CIT PET scans, bilateral antecubital veins were cannulated: one for blood sampling and for injection of ^18^F-FP-CIT, and the other for glucose or placebo infusions. The subjects were blinded and randomly assigned for either glucose or placebo infusions. Over 10 minutes, 300 mg/kg of glucose in a 50% solution, was administered. The placebo (normal saline) was also administered in the same speed and volume [19]. An intravenous bolus injection of ^18^F-FP-CIT was administered after the infusion of glucose or placebo. The emission data were acquired over 90 mins with 50 frames of progressively increasing durations (15 s × 8 frames, 30 s × 16 frames, 60 s × 10 frames, 240 s × 10 frames, and 300 s × 6 frames) using the Siemens Biograph 40 Truepoint PET/computed tomography (CT) (Siemens Healthcare, Knoxville, Tennessee, USA).

The dynamic PET data were collected in the 3-dimensional mode, with 148 slices with image sizes of 256 × 256 and pixel sizes of 1.3364 × 1.3364 mm^2^. These were reconstructed by filtered back projection using a Gaussian filter. The serum glucose level (mg/dL) and insulin level (μU/mL) were measured before and after the infusions of glucose and placebo. Serum glucose level was determined through an enzymatic reference method using hexokinase with the Glucose HK Gen.3 (Roche Diagnostics GmbH, Germany). Serum insulin level was determined through an electrochemiluminescence immunoassay method using Elecsys Insulin (Roche Diagnostics GmbH, Germany). For ^18^F-FDG PET scans, sixty minutes after injection of ^18^F-FDG, the emission data were acquired for 10 mins with Gemini TF PET/CT (Philips, Milpitas, CA, USA). The static PET data were collected in the 3-dimensional mode and reconstructed by the iterative ordered subset expectation maximization method.

### Image Analysis

Oxford-GSK-Imanova striatal atlas from FMRIB Software Library v5.0 (https://fsl.fmrib.ox.ac.uk/fsl) was applied, which is an atlas involving sub-striatal regions of ventral striatum (VST), caudate, and putamen segmented according to the anatomical structure, and manually delineated on the non-linear MNI 152 template [20]. For ^18^F-FP-CIT PET, an averaged image (0-10 min after injection) was created from dynamic PET frames and spatially normalized to ^15^O-Water PET template in statistical parametric mapping 5 (Wellcome Trust Centre for Neuroimaging, United Kingdom). DAT availability, expressed in terms of binding potential (BP_ND_), were measured by analyzing time-activity curve via the simplified reference tissue method [21] with a reference of cerebellum. For ^18^F-FDG PET, scans were normalized to ^15^O-Water PET template in statistical parametric mapping 5 (Wellcome Trust Centre for Neuroimaging, United Kingdom). The mean uptake of each striatal ROI was scaled to the mean of global cortical uptake of each individual, and defined as standardized uptake value ratio (SUVR). Image analysis was done with pmod version 3.6 (PMOD Technologies LLC, Zurich, Switzerland).

### Sweet taste questionnaire (STQ)

STQ consists of 12-item self-reporting to evaluate subjects’ reactions to eating sweets, craving for sweets and degree of control over eating sweet foods [22]. Scores for each item range between 1 (strongly disagree) and 7 (strongly agree) with higher scores indicating elevated sensitivity to mood altering effect of sweet foods and impaired control over eating sweets. STQ identifies two factors in the individual’s attitude towards sweet foods; STQ 1: sensitivity to mood altering effect of sweets, and STQ 2: impaired control over eating sweet foods.

### Statistical analysis

Normality was tested with Shapiro-Wilk test. Pearson correlation analysis was used to determine whether BMI, and serum glucose & insulin levels are associated with STQ 1, and STQ 2. Paired t-test was used to test the difference of serum glucose & insulin levels before, and after glucose loading, and the difference between glucose-loaded BP_ND_, and placebo-loaded BP_ND_ of ^18^F-FP-CIT PET. After logarithmic transformation of BP_ND_ and SUVR, the effects of STQ on BP_ND_ and SUVR were investigated using Bayesian hierarchical modelling with brms [23-25] that applies the Markov-Chain Monte Carlo sampling tools of RStan [26]. We created models separately with STQ 1, and STQ 2 as predictors, with BP_ND_ and SUVR as a dependent variable, adjusting for age. We added subject and ROI as random intercepts to allow BP_ND_ and SUVR to vary between subjects and ROIs. Bayesian models were estimated using four Markov chains, each of which had 4,000 iterations including 1,000 warm-ups, thus totaling 12,000 post-warmup samples. The sampling parameters were slightly modified to facilitate convergence (max treedepth = 20). Statistical analysis was carried out in R Statistical Software (The R Foundation for Statistical Computing).

## RESULTS

Thirty-five subjects (age 24.4±2.7 years, BMI 23.8±3.4 kg/m^2^) underwent two ^18^F-FP-CIT brain PET scans, and twenty-four underwent additional ^18^F-FDG brain PET scans. Mean ^18^F-FP-CIT and ^18^F-FDG brain PET scans are shown in Figure 1. STQ 1 (sensitivity to mood altering effect of sweets) ranged from 7 to 28 with the mean of 14.9±5.9. STQ 2 (impaired control over eating sweet foods) ranged from 5 to 19 with the mean of 9.1±4.3. Serum glucose level (before glucose loading: 85.2±9.4 mg/dL, after glucose loading: 117.6±48.3 mg/dL), and serum insulin level (before glucose loading: 7.0±4.4 μU/mL, after glucose loading: 19.7±21.3 μU/mL) were significantly increased after glucose loading (p<0.0001, p<0.0003). BMI was not correlated with either STQ 1 (r=-0.0868, p=0.6199) or STQ 2 (r=0.1027, p=0.5573). Serum glucose & insulin levels after glucose loading were not correlated with STQ 1 (r=-0.0066, p=0.97; r=-0.0063, p=0.9711), and STQ 2 (r=0.1276, p=0.4652; r=-0.0527, p=0.7637). From caudate, glucose-loaded DAT availability was significantly higher than placebo-loaded DAT availability (p=0.0495), and from putamen, glucose-loaded DAT availability showed the higher trend than placebo-loaded DAT availability (p=0.0761) (Figure 2).

**Figure 1.**
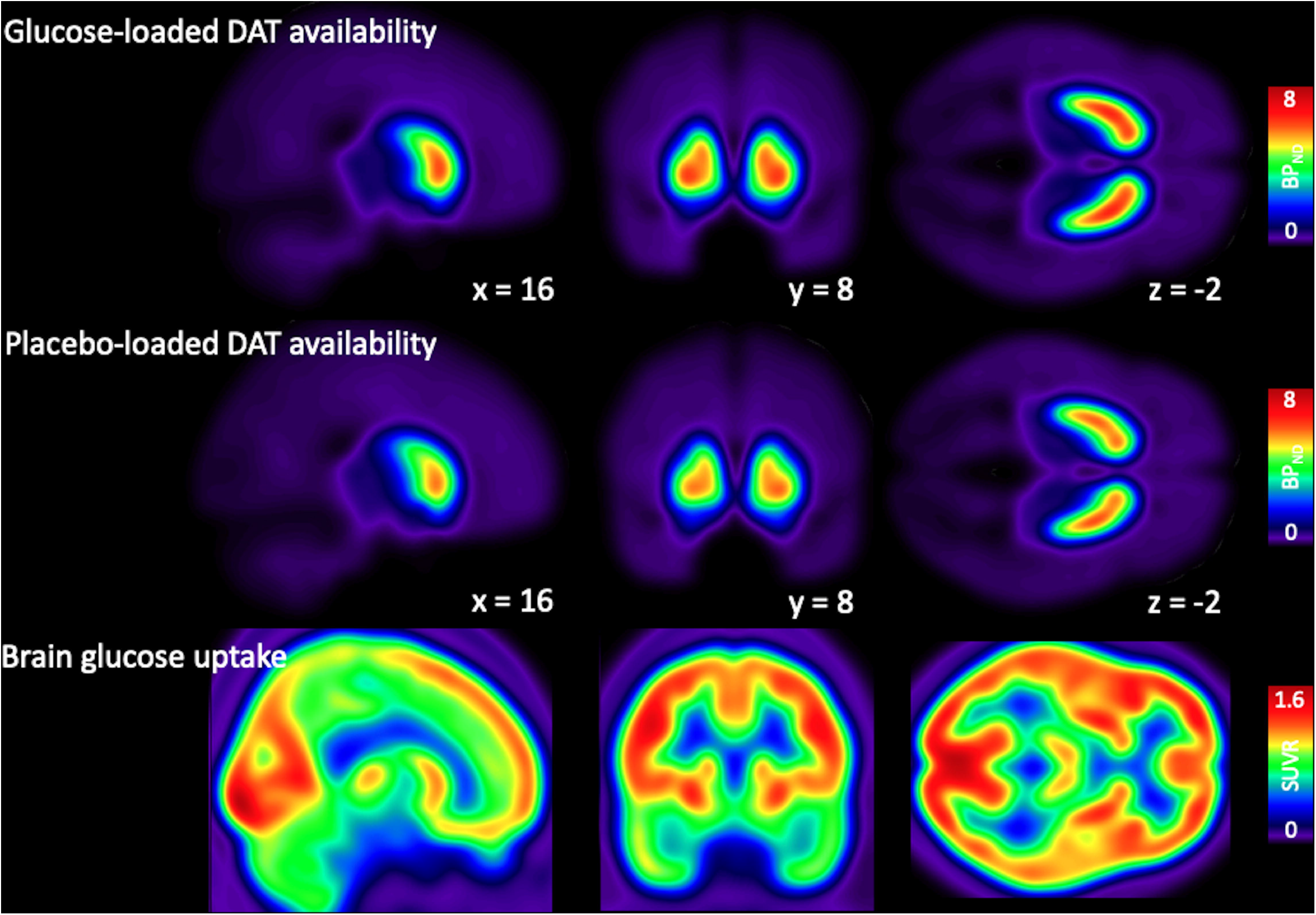
Mean ^18^F-FP-CIT (glucose-loaded, and placebo-loaded DAT availability) and ^18^F-FDG (brain glucose uptake) PET

**Figure 2.**
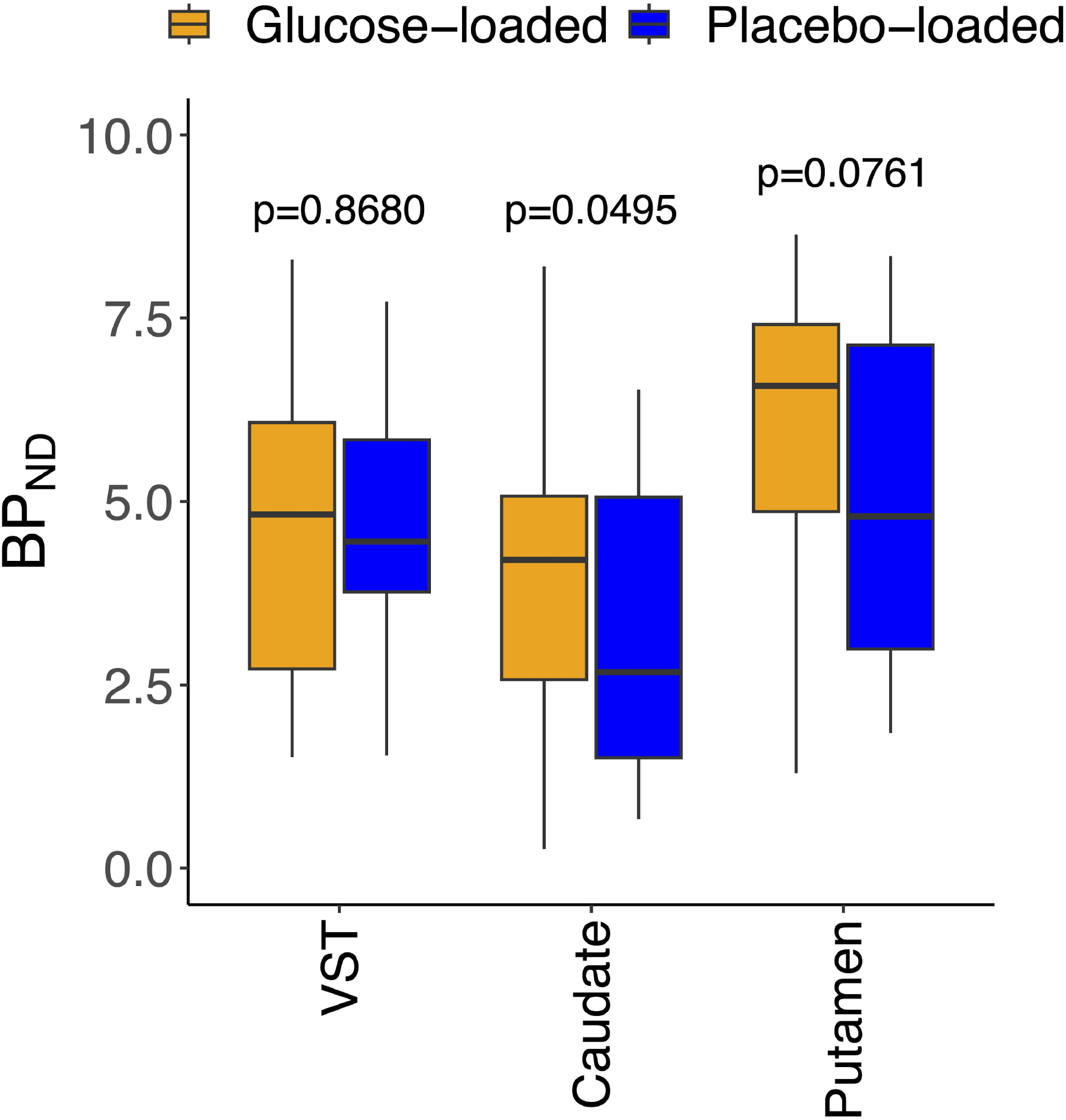
Paired t-test of glucose-loaded BP_ND_, and placebo-loaded BP_ND_ from ^18^F-FP-CIT PET

From Bayesian analysis, glucose-loaded DAT availability from VST, caudate, and putamen was positively associated with both STQ 1 (sensitivity to mood altering effect of sweets), and STQ 2 (impaired control over eating sweet foods). However, the effects of STQ 1 and STQ 2 on placebo-loaded DAT availability, and brain glucose uptake markedly overlapped with zero (Figure 3).

**Figure 3.**
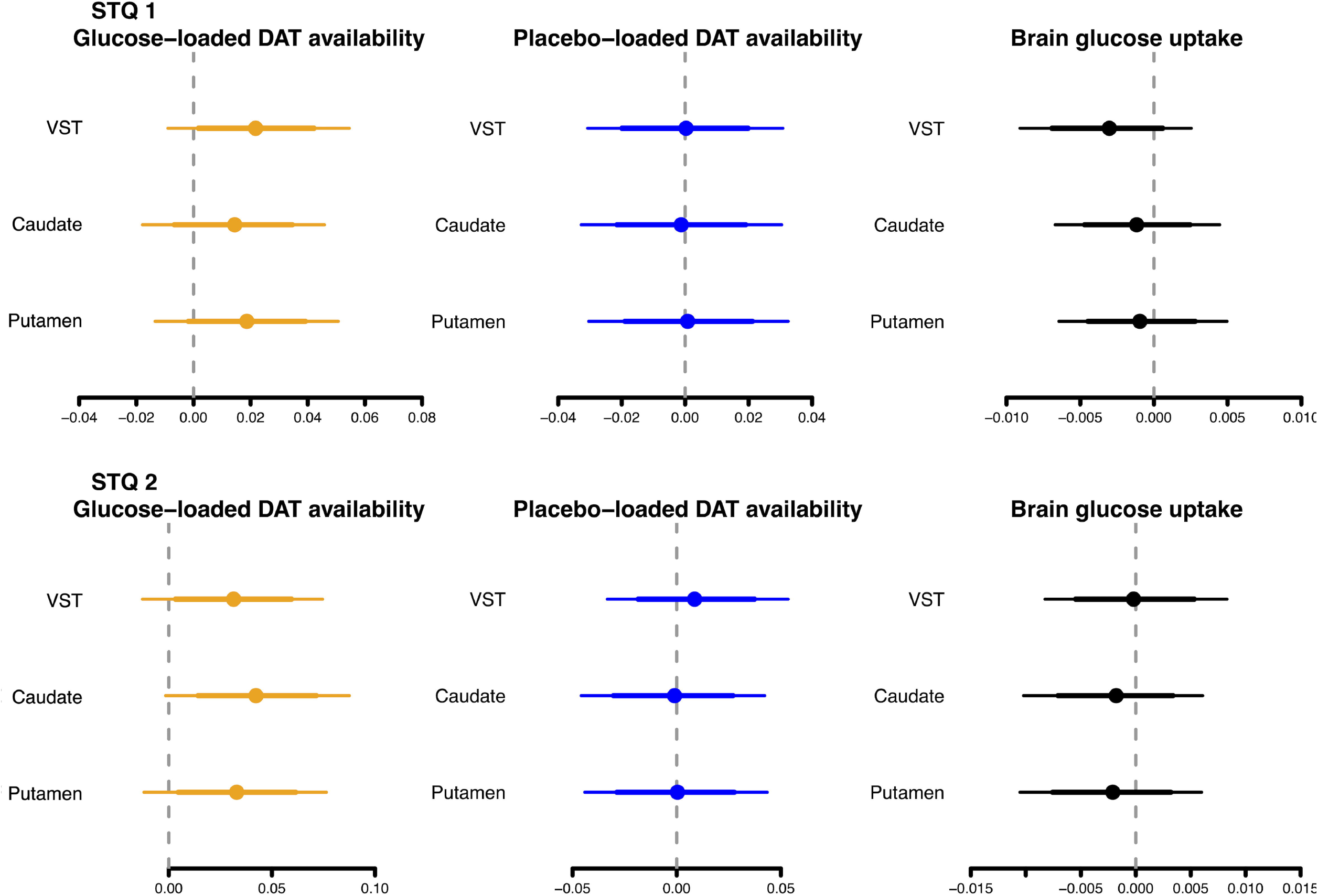
Posterior intervals of the regression coefficients of STQ 1, and STQ 2 for prediction of glucose-loaded DAT availability, placebo-loaded DAT availability, and brain glucose metabolism. The thick lines represent the 80% posterior intervals, the thin lines represent the 95% posterior intervals, and the circles represent the posterior means

## DISCUSSION

In this study, sensitivity to mood altering effect of sweets, and impaired control over eating sweet foods were associated with glucose-loaded DAT availability, not with placebo-loaded DAT availability or brain glucose metabolism. However, neither BMI, nor serum glucose & insulin levels were correlated with sensitivity to mood altering effect of sweets, and impaired control over eating sweet foods.

The brain plays a crucial role in controlling energy balance [7,27]. Eating behaviour is controlled by both homeostatic regulatory brain circuits and those involved in reward and motivation [28]. Dopamine is one of the neurotransmitters involved in eating behaviour, modulating the rewarding properties of food and the motivation and desire to consume food [9]. There are two major hypotheses regarding the role of dopamine in obesity. The first hypothesis, dopamine hyperresponsiveness, proposes that there is a hypersensitivity to rewards and increased behavioral salience toward food, resulting in the excessive intake of palatable foods [29,30]. The second hypothesis, reward deficit model, states that subjects who are insensitive to rewards overeat to increase their endogenous dopamine levels to a normal amount of pleasure [29,30]. However, it is not possible to measure the synaptic dopamine levels directly in the human brain. Therefore, PET with dopamine receptor (DR) radiopharmaceuticals has been used to understand the role of dopaminergic system in obesity. However, the recent studies showed that brain DR availability is not different between lean and overweight/obese subjects both from a meta-analysis [12], and a large PET cohort study [31]. In addition, it is difficult to interpret the relationships between DR availability measured from PET scans and the synaptic dopamine level. Low DR binding potential could represent 1) low density of existing DR, 2) low affinity to bind DR or 3) greater amount of endogenous dopamine which competes with DR radiopharmaceutical to bind DR [10,11].

DAT has not received as much attention in the field of eating behaviour until recently. DAT actively translocates the synaptic dopamine from the extracellular space into the presynatptic neuron, and is a major target for addictive drugs including cocaine, amphetamine, and methamphetamine [14]. From animal studies, insulin can act on striatal insulin receptors to enhance dopamine uptake by DAT through phosphatidylinositol 3-kinase signaling pathway, and downstream activation of Akt, which increases DAT surface expression [13], leading to clearance of synaptic dopamine [32]. Previously, the change of striatal DAT after glucose loading was reported by our group for the first time in humans [17]. We adopted the intravenous injection of glucose to reflect the biological aspects of food intake. Unlike DR, DAT is not affected by endogenous dopamine concentrations. Depletion of dopamine production by □-Methyl-*p*-tyrosine did not affect ^123^I-β-CIT binding in non-human primates [33]. In addition, dopamine release by haloperidol did not affect ^123^I-FP-CIT binding in rats [34], implying nearly irreversible binding of ^123^I-FP-CIT to DAT [35]. In this study, the change of BP_ND_ from ^18^F-FP-CIT PET scan was visualized in dorsal striatum, not in VST, after glucose loading. Therefore, this change of DAT availability after glucose loading might represent the non-hedonic response of striatal DAT by insulin signaling, regardless of synaptic dopamine concentration.

Interestingly, glucose-loaded DAT availability was positively associated with sensitivity to mood altering effect of sweets, and impaired control over eating sweet foods from STQ, suggesting that the subject with higher DAT availability after glucose loading is the more vulnerable to sweet foods. This may indicate that individuals with higher DAT availability after glucose loading experience a rapid clearance of synaptic dopamine after consuming sweet foods, potentially leading to a desire for additional sweet foods. Therefore, insulin signaling at dopamine terminals might contribute to the rewarding properties of food consumption [36]. However, there was no significant correlation between STQ, and BMI, serum glucose & insulin levels. Individuals with the preference for the stronger sweet solution have a stronger mood altering effect of sweet foods and an impaired control over eating sweets [22], and are linked with alcohol use disorder [37]. Human brain utilizes glucose as its main source of energy, thus, brain glucose uptake, assessed by ^18^F-FDG PET can be utilized for quantifying neuronal activity in the human brain [38]. However, the effects of STQ 1 and STQ 2 on placebo-loaded DAT availability, and brain glucose uptake markedly overlapped with zero, implying that the baseline DAT availability, and neuronal activity of striatum does not reflect the vulnerability to sweet foods.

There are several limitations to consider in this study. First, although 35 healthy subjects were included, the sample size was small. Second, only males were included in this study, thus these results may not directly generalize to females. Third, the BP_ND_s that were measured did not distinguish between the DAT density and affinity.

In conclusion, the change of striatal DAT availability after glucose loading is associated with the vulnerability to sweet foods. This may indicate that individuals with higher DAT availability after glucose loading experience a rapid clearance of synaptic dopamine after consuming sweet foods, potentially leading to a desire for additional sweet foods.

## Data Availability

All data produced in the present study are available upon reasonable request to the authors

## REFERENCES

1 Na SY, Myung SJ. [Obesity and colorectal cancer]. Korean J Gastroenterol. 2012;59(1):16–26.

2 Gukovsky I, Li N, Todoric J, Gukovskaya A, Karin M. Inflammation, autophagy, and obesity: common features in the pathogenesis of pancreatitis and pancreatic cancer. Gastroenterology. 2013;144(6):1199–209 e4.

3 Mijovic T, How J, Payne RJ. Obesity and thyroid cancer. Front Biosci (Schol Ed). 2011;3:555–64.

4 Alzahrani B, Iseli TJ, Hebbard LW. Non-viral causes of liver cancer: does obesity led inflammation play a role? Cancer Lett. 2014;345(2):223–9.

5 Gu W, Chen C, Zhao KN. Obesity-associated endometrial and cervical cancers. Front Biosci (Elite Ed). 2013;5:109–18.

6 Burke GL, Bertoni AG, Shea S, Tracy R, Watson KE, Blumenthal RS, et al. The impact of obesity on cardiovascular disease risk factors and subclinical vascular disease: the Multi-Ethnic Study of Atherosclerosis. Arch Intern Med. 2008;168(9):928–35.

7 Morton GJ, Meek TH, Schwartz MW. Neurobiology of food intake in health and disease. Nat Rev Neurosci. 2014;15(6):367–78.

8 Cameron JD, Goldfield GS, Cyr MJ, Doucet E. The effects of prolonged caloric restriction leading to weight-loss on food hedonics and reinforcement. Physiol Behav. 2008;94(3):474–80.

9 Bello NT, Hajnal A. Dopamine and binge eating behaviors. Pharmacol Biochem Behav. 2010;97(1):25–33.

10 Guo J, Simmons WK, Herscovitch P, Martin A, Hall KD. Striatal dopamine D2-like receptor correlation patterns with human obesity and opportunistic eating behavior. Mol Psychiatry. 2014;19(10):1078–84.

11 Janssen LK, Horstmann A. Molecular Imaging of Central Dopamine in Obesity: A Qualitative Review across Substrates and Radiotracers. Brain Sci. 2022;12(4).

12 Pak K, Nummenmaa L. Brain dopamine receptor system is not altered in obesity: Bayesian and frequentist meta-analyses. Hum Brain Mapp. 2023;44(18):6552–60.

13 Vaughan RA, Foster JD. Mechanisms of dopamine transporter regulation in normal and disease states. Trends Pharmacol Sci. 2013;34(9):489–96.

14 Thomsen G, Ziebell M, Jensen PS, da Cuhna-Bang S, Knudsen GM, Pinborg LH. No correlation between body mass index and striatal dopamine transporter availability in healthy volunteers using SPECT and [123I]PE2I. Obesity (Silver Spring). 2013;21(9):1803–6.

15 Nam SB, Kim K, Kim BS, Im HJ, Lee SH, Kim SJ, et al. The Effect of Obesity on the Availabilities of Dopamine and Serotonin Transporters. Sci Rep. 2018;8(1):4924.

16 van de Giessen E, Hesse S, Caan MW, Zientek F, Dickson JC, Tossici-Bolt L, et al. No association between striatal dopamine transporter binding and body mass index: a multi-center European study in healthy volunteers. Neuroimage. 2013;64:61–7.

17 Pak K, Seo S, Kim K, Lee MJ, Shin MJ, Suh S, et al. Striatal dopamine transporter changes after glucose loading in humans. Diabetes Obes Metab. 2020;22(1):116–22.

18 Pak K, Seo S, Lee MJ, Kim K, Suh S, Im HJ, et al. Striatal DAT availability does not change after supraphysiological glucose loading dose in humans. Endocr Connect. 2021;10(10):1266–72.

19 Haltia LT, Rinne JO, Merisaari H, Maguire RP, Savontaus E, Helin S, et al. Effects of intravenous glucose on dopaminergic function in the human brain in vivo. Synapse. 2007;61(9):748–56.

20 Tziortzi AC, Searle GE, Tzimopoulou S, Salinas C, Beaver JD, Jenkinson M, et al. Imaging dopamine receptors in humans with [11C]-(+)-PHNO: dissection of D3 signal and anatomy. Neuroimage. 2011;54(1):264–77.

21 Lammertsma AA, Hume SP. Simplified reference tissue model for PET receptor studies. Neuroimage. 1996;4(3 Pt 1):153-8.

22 Kampov-Polevoy AB, Alterman A, Khalitov E, Garbutt JC. Sweet preference predicts mood altering effect of and impaired control over eating sweet foods. Eat Behav. 2006;7(3):181–7.

23 Bürkner P-C. Bayesian Item Response Modeling in R with brms and Stan. Journal of Statistical Software. 2021;100(5):1–54.

24 Bürkner P-C. brms: an R package for Bayesian multilevel models using Stan. Journal of Statistical Software. 2017;80(1):1–28.

25 Bürkner P-C. Advanced Bayesian multilevel modeling with the R package brms. The R Journal. 2018;10(1):395–411.

26 Stan Development Team. RStan: the R interface to Stan. 2022. https://mc-stan.org/.

27 Pak K, Kim SJ, Kim IJ. Obesity and Brain Positron Emission Tomography. Nucl Med Mol Imaging. 2018;52(1):16–23.

28 Volkow ND, Wang GJ, Tomasi D, Baler RD. The addictive dimensionality of obesity. Biol Psychiatry. 2013;73(9):811–8.

29 Kessler RM, Zald DH, Ansari MS, Li R, Cowan RL. Changes in dopamine release and dopamine D2/3 receptor levels with the development of mild obesity. Synapse. 2014;68(7):317–20.

30 Verbeken S, Braet C, Lammertyn J, Goossens L, Moens E. How is reward sensitivity related to bodyweight in children? Appetite. 2012;58(2):478–83.

31 Malén T, Karjalainen T, Isojärvi J, Vehtari A, Bürkner P-C, Putkinen V, et al. Age and sex dependent variability of type 2 dopamine receptors in the human brain: A large-scale PET cohort. Neuroimage. 2022.

32 Mebel DM, Wong JC, Dong YJ, Borgland SL. Insulin in the ventral tegmental area reduces hedonic feeding and suppresses dopamine concentration via increased reuptake. Eur J Neurosci. 2012;36(3):2336–46.

33 Heinz A, Jones DW, Zajicek K, Gorey JG, Juckel G, Higley JD, et al. Depletion and restoration of endogenous monoamines affects beta-CIT binding to serotonin but not dopamine transporters in non-human primates. J Neural Transm Suppl. 2004(68):29–38.

34 Booij J, van Loon G, de Bruin K, Voorn P. Acute administration of haloperidol does not influence 123I-FP-CIT binding to the dopamine transporter. J Nucl Med. 2014;55(4):647–9.

35 Booij J, Hemelaar TG, Speelman JD, de Bruin K, Janssen AG, van Royen EA. One-day protocol for imaging of the nigrostriatal dopaminergic pathway in Parkinson’s disease by [123I]FPCIT SPECT. J Nucl Med. 1999;40(5):753–61.

36 Stouffer MA, Woods CA, Patel JC, Lee CR, Witkovsky P, Bao L, et al. Insulin enhances striatal dopamine release by activating cholinergic interneurons and thereby signals reward. Nat Commun. 2015;6:8543.

37 Braun TD, Kunicki ZJ, Blevins CE, Stein MD, Marsh E, Feltus S, et al. Prospective Associations between Attitudes toward Sweet Foods, Sugar Consumption, and Cravings for Alcohol and Sweets in Early Recovery from Alcohol Use Disorders. Alcohol Treat Q. 2021;39(3):269–81.

38 de Leon MJ, Convit A, Wolf OT, Tarshish CY, DeSanti S, Rusinek H, et al. Prediction of cognitive decline in normal elderly subjects with 2-[(18)F]fluoro-2-deoxy-D-glucose/poitron-emission tomography (FDG/PET). Proc Natl Acad Sci U S A. 2001;98(19):10966–71.

